# Integrating genomic data into test-negative designs for estimating lineage-specific COVID-19 vaccine effectiveness

**DOI:** 10.1101/2025.07.10.25331291

**Authors:** Kevin C. Ma, Diya Surie, Natalie Dean, Clinton R. Paden, Natalie J. Thornburg, Fatimah S. Dawood

**Author notes:** Correspondence: Kevin C. Ma, Centers for Disease Control and Prevention, 1600 Clifton Road, NE, Atlanta, Georgia, USA.

## Abstract

**Background:** SARS-CoV-2 lineage-specific COVID-19 vaccine effectiveness (VE) studies can inform decision-making on whether vaccine composition updates are needed to maintain effectiveness against severe disease as SARS-CoV-2 continues to evolve. Lineage assignment methods in VE test-negative design (TND) studies include sequence-based (whole-genome sequencing), proxy-based (e.g., S-gene target failure during polymerase chain reaction), and period-based (variant predominance time periods) approaches.

**Methods:** We first summarize benefits, challenges (including cost and timeliness), and methodologic considerations related to estimating lineage-specific COVID-19 VE against emerging variants using these different assignment approaches. We then use a deterministic model to illustrate the potential impact of lineage misclassification error on VE estimates in a TND using period-based versus sequence-based lineage assignment across different variant emergence scenarios.

**Results:** Our model suggests period-based analyses may underestimate differences in VE between two lineages due to lineage misclassification error in certain variant circulation scenarios. This effect is most evident during prolonged variant co-circulation or in early time periods following new variant takeover. Using higher predominance thresholds can reduce VE estimate bias in period-based analyses but at the expense of sample size, reducing precision or outright precluding estimation under some scenarios. Period-based VE analyses should therefore consider including sensitivity analyses to characterize robustness of VE estimates to different predominance thresholds.

**Conclusions:** TND studies using sequence-, proxy-, and period-based lineage assignment have contributed substantially towards understanding SARS-CoV-2 variant-mediated vaccine escape, but biases that can affect each study design vary. Our results identify analytic considerations for robust estimation and suggest principles that may translate to other pathogens that undergo continuous antigenic drift.

**Summary:** Whole-genome sequencing, predominance time periods, and genetic proxies are common lineage assignment approaches used to estimate variant-specific COVID-19 vaccine effectiveness. These approaches have varied strengths, costs, and biases that may affect the accuracy and precision of vaccine effectiveness estimates.

## Introduction

The continued emergence of SARS-CoV-2 lineages with increased immune evasion from COVID-19 vaccines and therapeutics presents a public health challenge. The earliest signals of immune escape are often obtained through laboratory characterization of neutralizing antibody titers against emerging variants using sera from vaccinated individuals [1–6]. Timely epidemiologic estimates of COVID-19 vaccine effectiveness (VE) against SARS-CoV-2 variants provide important real-world validation of serologic signals to guide public health risk assessments and decisions about updating COVID-19 vaccine composition [7].

The test-negative design (TND) has been increasingly applied for estimating overall and lineage-specific COVID-19 VE, but methods vary for integrating viral genetic data and addressing confounding and other biases [8–15]. A key consideration is the method used to ascertain and classify SARS-CoV-2 lineages. Common methods for lineage assignment include 1) sequence-based methods using whole-genome sequencing (WGS) of SARS-CoV-2 specimens; 2) proxy-based methods relying on a genetic marker associated with a lineage of interest, such as the spike protein residue 69–70 deletion resulting in S-gene polymerase chain reaction (PCR) amplification failure [16,17]; and 3) time period-based assignment using variant predominance periods. Importantly, these approaches have varied strengths, costs, timeliness, and susceptibility to biases that may affect the accuracy and precision of lineage-specific VE estimates.

Here, we highlight benefits, challenges, and methodologic considerations for different lineage assignment approaches when estimating COVID-19 VE against emerging and co-circulating SARS-CoV-2 variants. We use a simple deterministic model to illustrate the potential impact of lineage misclassification error on VE estimates based on period-based assignment across different variant emergence scenarios.

### Lineage assignment approaches in TND VE studies

#### Sequence-based

Sequence-based approaches rely on whole-genome sequencing (WGS) of SARS-CoV-2 cases for assigning lineage, which remains the gold standard for variant identification. Sequence-based methods were used throughout the pandemic to assess VE as new variants emerged [18–24]. For example, Kirsebom et al. assessed VE of XBB.1.5 and BA.4/5 boosters given as part of the fall 2023 program in the UK against XBB, EG.5.1, and JN.1 lineages using WGS. The authors found evidence for lower VE against JN.1 lineages, which emerged in late 2023 and became predominant in January 2024 [21], mirroring findings from a multi-site cohort in the US which also used WGS to assess VE against XBB and JN lineages [13]. Unlike proxy- or period-based methods, sequence-based approaches have the advantage of full genomic resolution and could potentially also be used to assess VE by specific spike protein substitutions [25–27] or by genetic distance from the COVID-19 vaccine strain [28,29]. Methods to characterize how vaccine efficacy varies by infecting pathogen strain have been referred to as sieve analyses and have been conducted for HIV, malaria, and SARS-CoV-2 vaccines in randomized clinical trials [30–33], but are not yet routinely applied in observational vaccine studies.

Sequence-based lineage assignment using WGS requires active sample collection or salvaging of stored clinical samples (potentially including rapid antigen tests [34]) and efficient specimen shipping, processing, and sequencing to produce timely VE estimates. Completeness of sequencing results also depends on adequate respiratory specimen quality; specimens with high PCR cycle threshold values often contain insufficient viral material for sequencing. Sequence-based lineage assignment may introduce selection biases, as specimens that cannot be sequenced due to low viral load are excluded and viral load may correlate with variant, disease severity, or vaccination status [35–38]. Sensitivity analyses can be conducted comparing overall VE with VE among patients with sequenced specimens to assess the degree of bias [30]. Additionally, mixed infection with multiple SARS-CoV-2 strains can rarely occur, and lineage identification approaches need to distinguish between sequencing artifacts, true mixed infections, or novel variants [39]. Lastly, attaining adequate sample sizes for sufficient statistical power in VE studies is perhaps the greatest challenge for studies based on WGS compared to other assignment approaches due to cost, equipment, and personnel requirements for sequencing. This may preclude precise estimates of VE stratifying simultaneously by lineage and other covariates including age, immune status, and time since vaccination.

#### Proxy-based

In the absence of WGS capacity, real-time reverse transcription PCR against select amplification targets can sometimes be used as a proxy for genetic lineage (i.e. proxy-based lineage assignment). For SARS-CoV-2, a commonly used proxy is the TaqPath COVID-19 multiplexed diagnostic test, which amplifies the *S*, *ORF1ab*, and *N* genes of the viral genome [16]. Failure of *S* gene amplification can occur due to a deletion at spike protein residues 69 and 70. This mutation was first observed with the Alpha variant [40,41], and has since alternated in occurrence with the emergence of some subsequent variants, including Delta, Omicron, and various Omicron descendants [17,42]. Targeted RT-PCRs for specific spike mutations have also been previously used to detect known variants of concern [43,44], and further work is needed to develop new scalable and accurate proxies [45,46].

The use of S-gene target failure (SGTF) or presence (SGTP) as a lineage marker can enable rapid estimation of lineage-specific VE [9,47–49]. Tseng et al. used SGTP/F in a large integrated health network to estimate effectiveness of mRNA-1273 boosters against Delta and Omicron variant infection, respectively, in December 2021 [50]. Similarly, Link-Gelles et al. used SGTP/SGTF in a nationwide pharmacy testing network to conduct early estimates of VE against symptomatic XBB lineage infection in 2023 [8,9] and JN lineage infection in 2024 [49]. Use of SGTP/F to define lineages can be more cost-effective and higher-throughput than WGS because SGTP/F relies on existing diagnostic RT-PCR laboratory infrastructure, and results are typically available more quickly [17,42,51]. However, SGTP/F cannot discriminate between specific mutations or other sub-lineage variation that may confer immune escape.

One of the main challenges for proxy-based approaches is that sensitivity and positive predictive value for emerging variants of interest must be continuously assessed and validated using WGS. The sensitivity of SGTP/F for differentiating lineages has varied, and can be increased by including “partial” SGTF samples where amplification of the S-gene occurs but is reduced compared to other genomic targets [42,52]. The positive predictive value of SGTP/F can be influenced by prevalence, especially as a new variant is beginning to emerge. In December 2022, as the XBB.1.5 variant began to spread in the US, positive predictive value of SGTP was relatively low with XBB or XBB.1.5 constituting approximately 60% of SGTP specimens and the remainder being other BA.2 sublineages [9,17]. Andeweg et al. restricted to time periods when SGTP/F had >85% positive predictive value for variants of interest to address this issue [47], but this could delay VE estimation for emerging variants. Further assessments are needed to understand how this type of misclassification could influence VE estimates and to develop approaches to adjust for misclassification, such as regression calibration [53].

#### Time period-based

Period-based VE analyses approximate lineage assignment by assuming that all case-patients enrolled during a specified time period are infected by the predominant circulating SARS-CoV-2 variant. The period is defined using a prevalence threshold for the variants of interest, and thresholds used have varied considerably across studies: 50% is common [10,19,54–57], but higher thresholds such as 60% [11,58–60], 70% [61], 75% [62], 80% [12,63–65], and 90% [66] have also been used. A key advantage of this assignment approach is that it enables estimation of lineage-specific VE from platforms without individual-level WGS or PCR laboratory data, potentially enabling larger sample sizes and representativeness. For instance, Schrag et al. used a period-based approach in a large multisite electronic health record network to characterize mRNA VE against medically attended illness during Delta versus Omicron periods, with simultaneous stratifications by pregnancy status, number of doses, and days since dose receipt [57]. Period-based approaches cannot resolve variants during prolonged periods of co-circulation, such as during mid-2024 onwards, when multiple JN.1 descendants (e.g. KP.2, KP.3, LB.1, KP.3.1.1, XEC, etc.) emerged and co-circulated without establishing sustained predominance [67]. Period-based approaches may also face difficulty disentangling the effect of lineage on VE in the presence of time-varying confounders, such as waning VE from antibody level decreases following increasing time since vaccination. Extensions of period-based analyses to incorporate variant proportion as a continuous covariate may expand utility to a broader range of co-circulation scenarios [68].

Variant prevalence data for defining predominance periods can be derived using sequencing data from clinical specimens collected from the study population [54,55] or from external data sources. External sources include global sequence repositories [69] and surveillance programs such as the US CDC’s national genomic surveillance platform, which aggregated genomic data from CDC-contracted laboratories, the National Strain Surveillance program, and public sequence data repositories [70]. Sequencing data availability and capacity varies substantially globally [71,72], but if external data sources are available, period-based analyses likely offer cost-savings over sequence- or proxy-based approaches as it does not require collection, storage, and processing of SARS-CoV-2 clinical specimens. Robust period-based VE assessments therefore rely on sustained support for genomic surveillance programs, and decreasing counts of published sequences in some national surveillance platforms present a challenge for timely estimation of VE [73,74]. Additionally, due to geographic heterogeneity in variant circulation patterns, local variant prevalence at TND enrollment sites may not match external data sources. For instance, in the US, the XBB.1.5 lineage first appeared in the Northeast in late 2022 and attained predominance several weeks later on the West Coast [70]. Defining site-specific predominance periods in multicenter analyses may better account for regional differences in variant circulation [54,57], provided that sufficient regional sequencing data are available.

#### Other study designs

Some investigators have used a hierarchy of data sources to classify variants [20,24,55,63,75]. For instance, Adams et al. and Lauring et al. conducted analyses in a multi-site TND network using periods defined by both WGS of case-patients and a 50% prevalence threshold. Cases with a sequenced discordant lineage in a given period were excluded, reducing misclassification error where possible [20,55]. Tartof et al. used a hierarchy of approaches beginning with WGS when available, SGTF, and finally period-based classification (using an 80% or higher threshold), maximizing the number of included case-patients [63,75].

Additionally, some studies lack test-negative controls but can still provide evidence to evaluate serologic signals of immune escape from updated COVID-19 vaccines. In these case-only study designs, the adjusted odds of receipt of updated COVID-19 vaccination are compared between two lineage groups of interest [76–78], which can provide a relative degree of vaccine immune escape. Moustsen-Helms et al. used this approach for JN.1 versus XBB lineages to provide early evidence for increased immune escape by JN.1 lineages [77] that later TND assessments corroborated [79]. Both case-only study designs and traditional TNDs can also support early detections of changes in COVID-19 clinical severity as new variants emerge [13,18,77,78,80].

### Potential impact of lineage misclassification errors

#### Methods

We conducted a modeling study to assess the potential impact of lineage misclassification errors using period-based versus sequence-based approaches for estimating VE. We used empirical US COVID-19 incidence and genomic surveillance data to inform model structure and parameters by estimating COVID-19 cases attributable to variants circulating in the US during July 2021 to September 2024 (Supplementary Methods). We found that most variant epidemic curves were symmetric with variable overlap between successive variants and an average standard deviation of approximately 6 weeks (Supplementary Figure 1). To mirror these dynamics, we used a simple deterministic model for two co-circulating variants assuming normally-distributed epidemic curves with the peak of variant 2 circulation occurring a range of 2 to 30 weeks after the peak of variant 1, allowing parametric control over the extent of co-circulation (Supplementary Methods). We assessed other strategies for generating co-circulation patterns, such as varying the standard deviation of the variant 2 epidemic curve while holding peak timing constant (Supplementary Figure 2), but these approaches did not produce prolonged (>30 weeks) co-circulation.

Time-invariant VE against COVID-19 hospitalization was assumed to be 50% against variant 1 and 20% against variant 2, representing approximate differences in VE observed empirically against recent vaccine strain-matched and divergent variants, respectively [13,21,59,65]. These estimates of COVID-19 VE represent additional protection beyond existing levels conferred by prior vaccination or infection, which has been increasingly difficult to ascertain. We explored the impact of time-varying VE using the piecewise function approach of Lin et al. [81]. In this model, VE increases from 𝜏 = 0 to a peak at 𝜏 = 4 weeks of 50% against variant 1 and 20% against variant 2; VE then subsequently wanes to 0% to represent natural waning of immunologic protection over time (Supplementary Methods, Supplementary Figure 3). These parameters represent illustrative scenarios and are not meant to be best estimates of current rates of waning of COVID-19 VE.

Vaccine coverage was assumed to increase linearly from the start of the season before plateauing at 20% [82]. Incidence of non-SARS-CoV-2 acute respiratory infection resulting in hospitalization was assumed to be constant. Using a TND framework, we estimated lineage-specific VE using logistic regression adjusting for calendar time (categorical week), comparing VE estimate bias and precision when assigning lineages with sequence-based and period-based approaches across different predominance thresholds.

#### Results

By varying the peak of variant 2 circulation relative to variant 1, we generated a range of variant co-circulation scenarios. Setting variant 2 circulation to peak two weeks after variant 1 yielded prolonged co-circulation (37/39 weeks of circulation where both variant 1 and 2 were >10% prevalence) (Figure 1A). With increasing delay in the start of variant 2 circulation, variant epidemic curves had reduced overlap and variant 1 predominance transitioned more rapidly to variant 2 predominance (Figure 1C–F, Supplementary Figure 4). These scenarios correspond to the range of variant dynamics observed since 2021 in the US, such as prolonged co-circulation of multiple XBB descendant lineages in summer and fall 2023 followed by rapid takeover of XBB by JN.1 by January 2024 [73].

**Figure 1.**
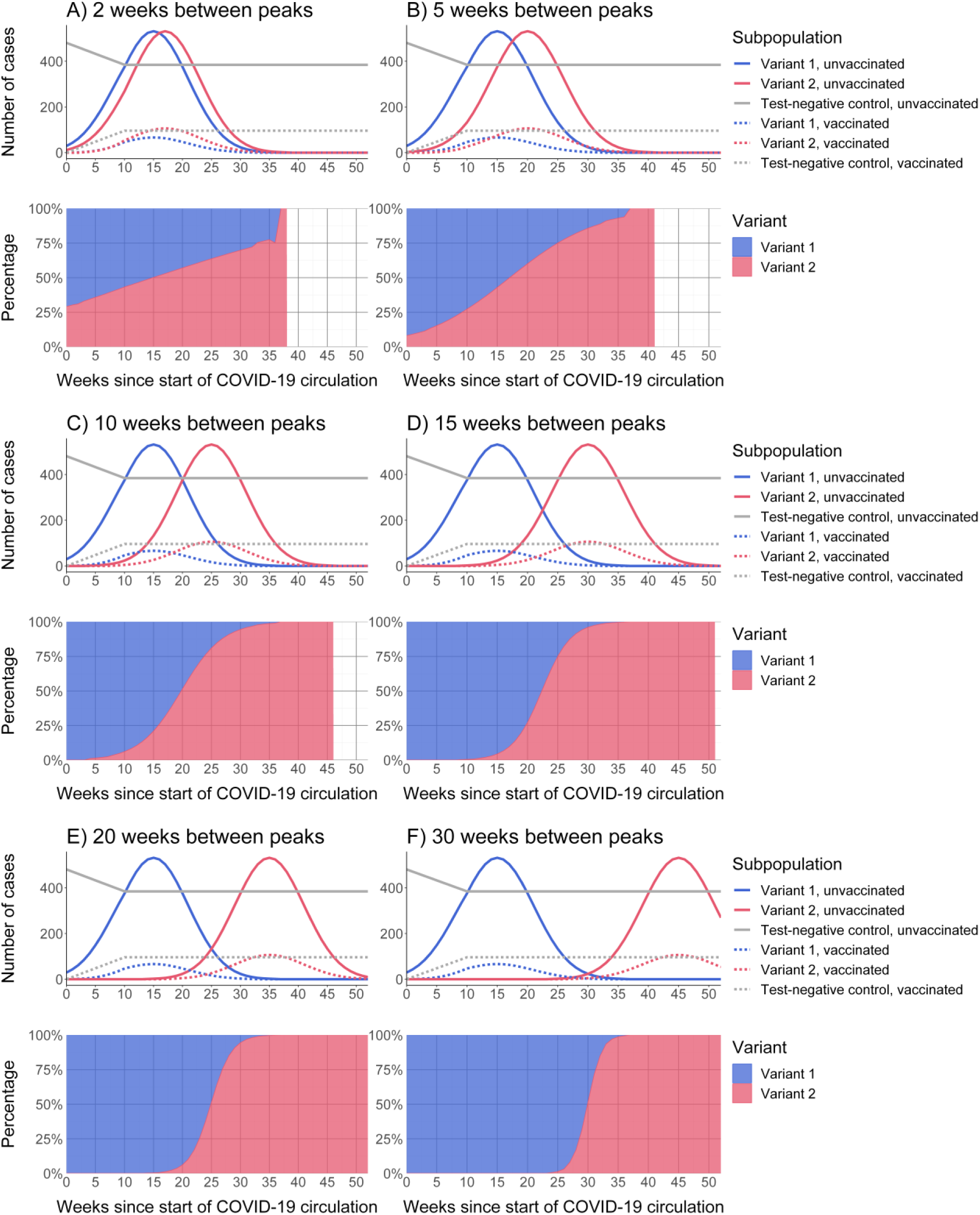
**Counts of simulated cases and controls by variant and vaccination status (top inset) and proportions of variants (bottom inset) over time for a range of co-circulation scenarios (A-F).** Variant co-circulation was modeled using symmetric normally-distributed epidemic curves, with the peak of variant 2 circulation occurring a range of 2 to 30 weeks after variant 1. Time-invariant VE was assumed to be 50% against variant 1 and 20% against variant 2, representing approximate differences in VE observed empirically against recent strain-matched and divergent variants, respectively. Vaccine coverage was assumed to increase linearly from the start of the season before plateauing at 20%, derived from empirical trends in vaccination coverage 2023–24 and 2024–25 in the US. *Alt text:* Multiple subfigures displaying line graphs of counts of simulated cases and controls by variant and vaccination status and area plots of proportions of variants over time for a range of co-circulation scenarios.

We assessed how period-based lineage-specific VE analyses using a 50% predominance threshold would perform under different variant dynamics scenarios. With prolonged co-circulation, a period-based analysis using a 50% threshold exhibited notable VE estimate bias (>10 percentage points for both variants) and underestimated the difference in VE between lineages (Figure 2A, Supplementary Table 1). As the duration of co-circulation decreased (i.e., sharper transition between variant periods), cases with misclassified lineage represented an increasingly smaller share of data and the bias in VE estimates decreased for both variants.

**Figure 2.**
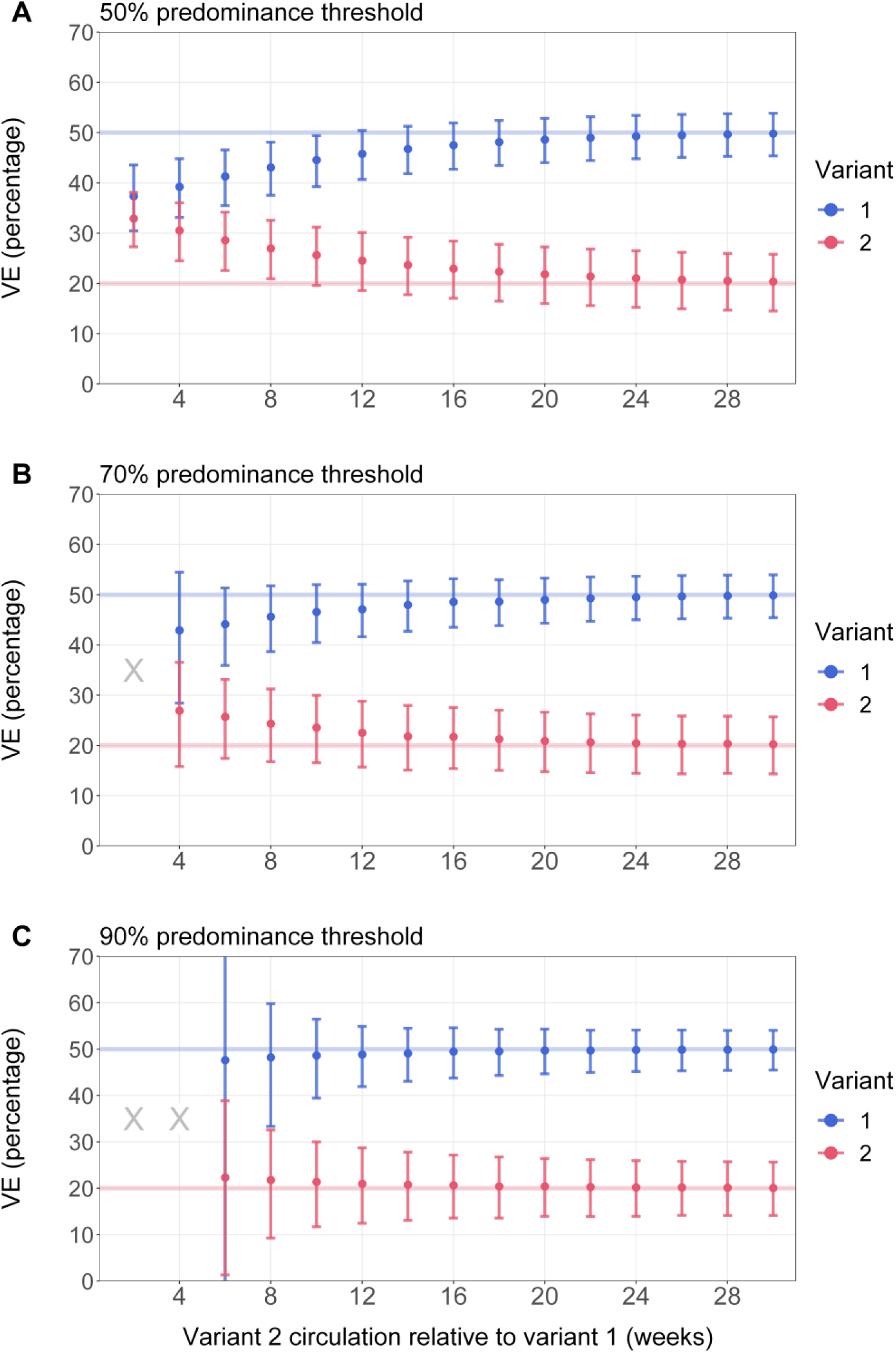
**Point estimates and 95% confidence intervals for period-based vaccine effectiveness estimates across different variant co-circulation scenarios and predominance thresholds.** X = scenarios during which a predominance threshold was not met or the number of COVID-19 cases was <100 for either variant and VE could not be estimated. Variant co-circulation was modeled using symmetric normally-distributed epidemic curves, with the peak of variant 2 circulation occurring a range of 2 to 30 weeks after variant 1. Time-invariant VE was assumed to be 50% against variant 1 and 20% against variant 2, representing approximate differences in VE observed empirically against recent strain-matched and divergent variants, respectively. Vaccine coverage was assumed to increase linearly from the start of the season before plateauing at 20%, derived from empirical trends in vaccination coverage 2023–24 and 2024–25 in the US. Using a TND framework, we estimated lineage-specific VE using logistic regression adjusting for calendar time, and we estimated lineage-specific VE using a period-based analysis with different predominance thresholds. *Alt text:* Point estimates and 95% confidence intervals depicted using error bars for variant 1 and 2 (colors) for period-based vaccine effectiveness estimates. Estimates are shown across different variant co-circulation scenarios along the x-axis and predominance thresholds (50%, 70%, 90%) in separate subfigures.

Because lineage misclassification during variant transition periods appeared to drive estimate bias, one solution was to increase the predominance threshold and effectively exclude weeks in which a substantial fraction of cases is likely to be misclassified. During periods in which estimation was possible, increasing the threshold to 70% (Figure 2B) or 90% (Figure 2C) reduced point estimate bias. However, by excluding samples, standard errors increased and confidence intervals widened (Supplementary Table 1). Additionally, in some scenarios with high co-circulation, predominance periods could not be defined or sample sizes were insufficient (number of COVID-19 cases for either variant <100) to estimate VE, highlighting a tradeoff between VE estimate precision and bias for period-based analyses (Figure 2B and 2C). When the modeled difference in VE between the two variants was reduced from 30% to 15% in a sensitivity analysis, the impact of this variant misclassification bias on absolute VE point estimate error was smaller, though relative trends were still similar (Supplementary Figure 5). Increasing the population-wide vaccination coverage level enhanced precision of estimates but did not substantially affect point estimates or the nature of bias observed (Supplementary Figure 6).

These findings suggested lineage misclassification error may be mitigated in scenarios in which a new variant quickly overtakes an old one. Such scenarios are often of epidemiologic and scientific interest as they suggest a fitness advantage for the new variant, but rapid estimation of VE as variant 2 is emerging is critical for early assessments of risk and public health impact [83]. Therefore, we modified the scenario where variant takeover was rapid (Figure 1E) by estimating VE weekly following variant 2 reaching >50% prevalence, mirroring challenges of limited data availability early in emerging variant growth trajectories. Using a sequence-based approach, estimates were unbiased and initially imprecise, but increasingly gained precision as data accumulated (Figure 3A, Supplementary Table 2). In contrast, a 50% period-based analysis substantially underestimated the difference in VE between lineages early, though both precision and bias also improved with accumulation of data (Figure 3B). Use of more stringent predominance thresholds reduced bias in the point estimate, but precision decreased and estimation was delayed until the prevalence of variant 2 reached the higher thresholds of 70% and 90%, respectively (Figure 3C, Figure 3D, Supplementary Table 2). Allowing VE to wane by time since vaccination yielded similar findings: VE estimation bias was highest for the 50% period-based analysis and lowest for the 90% period-based analysis during weeks when estimation was possible (Supplementary Figure 7).

**Figure 3.**
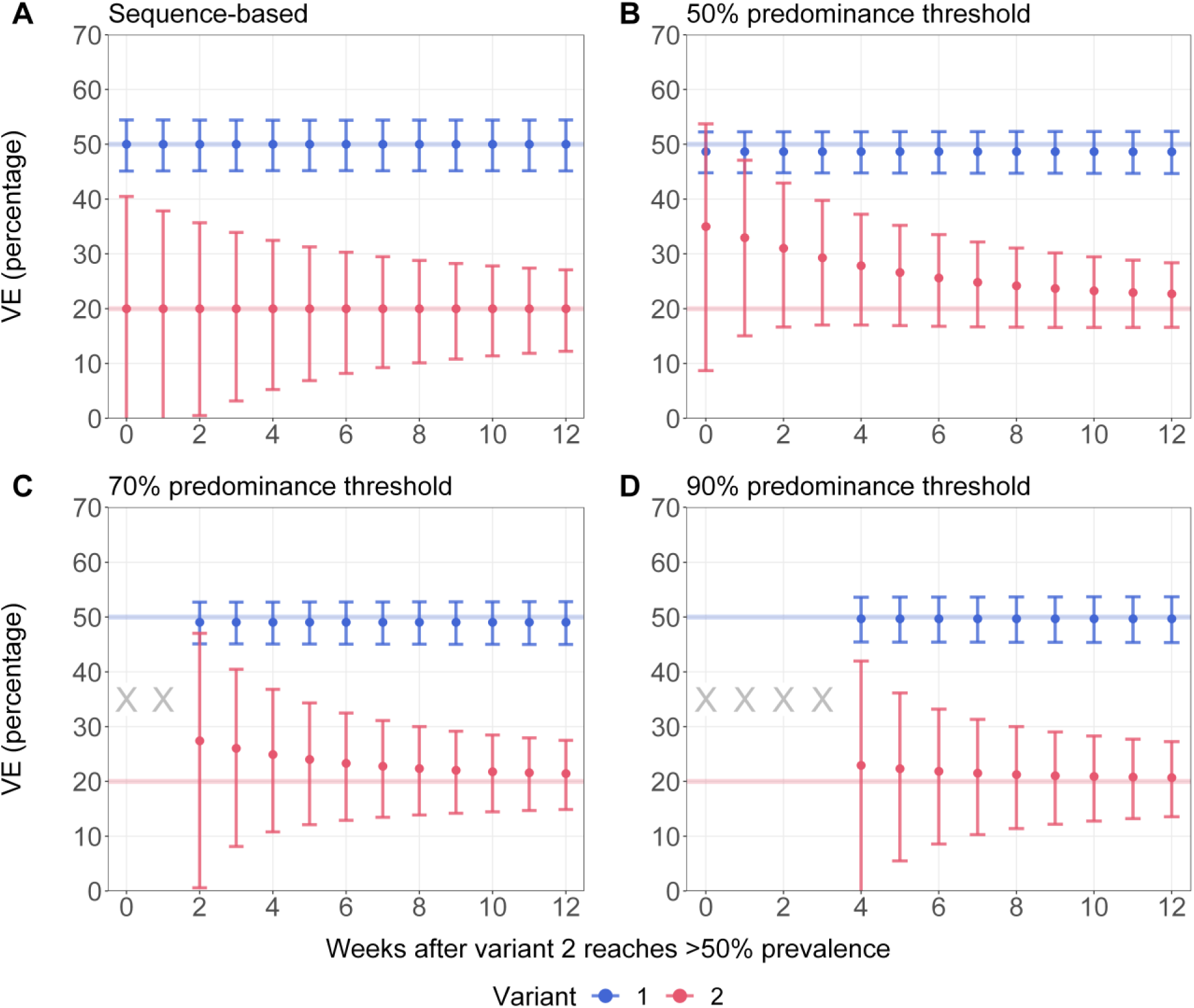
**Point estimates and 95% confidence intervals for (A) sequence-based and (B-D) period-based vaccine effectiveness estimates across different predominance thresholds by week after variant 2 reaches 50% prevalence.** X = scenarios during which a predominance threshold was not met or the number of COVID-19 cases was <100 for either variant and VE could not be estimated. Variant co-circulation was modeled using symmetric normally-distributed epidemic curves, with the peak of variant 2 circulation occurring a range of 2 to 30 weeks after variant 1. Time-invariant VE was assumed to be 50% against variant 1 and 20% against variant 2, representing approximate differences in VE observed empirically against recent strain-matched and divergent variants, respectively. Vaccine coverage was assumed to increase linearly from the start of the season before plateauing at 20%, derived from empirical trends in vaccination coverage 2023–24 and 2024–25 in the US. Using a TND framework, we estimated lineage-specific VE using logistic regression adjusting for calendar time, and we estimated lineage-specific VE using a period-based analysis with different predominance thresholds. *Alt text:* Point estimates and 95% confidence intervals depicted using error bars for variant 1 and 2 (colors) for period-based vaccine effectiveness estimates. Estimates are shown by number of weeks after variant 2 reaches >50% prevalence along the x-axis and lineage assignment method (sequence-based, or period-based using 50%, 70%, 90% thresholds) in separate subfigures.

## Discussion

Through a deterministic simulation, we highlight how period-based analyses may underestimate differences in VE between two lineages due to lineage misclassification error in certain scenarios. This issue was most pronounced during prolonged variant co-circulation or in early time periods following rapid variant takeover. Using higher predominance thresholds might reduce VE estimate bias at the expense of sample size, reducing precision or precluding estimation under some scenarios. Period-based VE analyses should therefore consider including sensitivity analyses to characterize robustness of VE estimates to different predominance thresholds. Sequence-based analyses do not face this tradeoff but can introduce selection bias (not included in this model) as not all case-patients will have successful specimen sequencing results [38].

TND studies using sequence-, proxy-, and period-based lineage assignment have all contributed substantially towards our understanding of SARS-CoV-2 variant-mediated vaccine escape, but biases that can affect these different study designs warrant evaluation. Under real-world settings, observed decreases in VE may be attributable to multiple factors beyond lineage replacement, and disentangling the impact of specific factors such as natural or “spurious” waning versus variant replacement can be challenging [8,84–86]. Findings from this analysis are meant to illustrate important considerations for estimating lineage-specific VE rather than provide definitive estimates of the magnitude of bias. Empirical comparisons of lineage assignment approaches, ideally within a single study platform, remain important to inform interpretation of VE estimates and inform methodological best practices [19].

This model is subject to several limitations. First, we did not model biases that affect sequence-based approaches, including delays in generating sequencing data and selection bias for specimens with successful sequencing. Second, to focus on the effect of lineage misclassification error, we used a basic infection model that did not account for risk heterogeneity, prior SARS-CoV-2 infection or COVID-19 vaccination, or depletion of susceptible individuals [85]. In addition, we did not model other general sources of bias in VE TNDs like confounding or misclassification of vaccination status, which have been reviewed and evaluated elsewhere [14,15,87–93]. Third, we limited assessment to two distinct lineages and did not model cross-reactive immunity between variants or account for stochastic effects that may cause variant extinction. Finally, based on empirical trends in the US, we assumed fairly rapid uptake of COVID-19 vaccine at the start of the circulation period with minimal increases in uptake after the initial weeks.

Lineage-specific VE studies can inform decision-making about whether updates to COVID-19 vaccine composition are needed to maintain protection against severe disease as SARS-CoV-2 continues to evolve [87]. Sustained SARS-CoV-2 WGS surveillance is critical to support lineage-specific VE studies, whether directly integrated into a TND through a sequence-based analysis or through epidemiologic surveillance sequencing to guide determination of predominance periods for period-based studies. Our results highlight how period-based VE analyses can benefit from assessing VE using different predominance thresholds, and add to the growing number of analytic considerations for TNDs [91], including accounting for prior infection [85,94], correlated vaccination behaviors [95,96], calendar time [13,84,97,98], and other potential confounders and biases [87,89–91,99]. Findings from this analysis can support high-quality national and global COVID-19 VE assessment strategies and suggest principles that may translate to other pathogens that undergo continuous antigenic drift, such as influenza viruses and rotavirus [100–102].

## Funding

This research received no external funding.

## Conflicts of interest

NED reports research grants from CDC, NIH, and the Gates Foundation; consulting fees from Janssen; support for attending meetings and/or travel from CEPI; and participation on an advisory or data safety monitoring board for the International Vaccine Institute. All other authors declare no competing interests.

## Disclaimer

The findings and conclusions in this report are those of the authors and do not necessarily represent the official position of the Centers for Disease Control and Prevention (CDC).

## Data Availability

Simulation results are available in supplementary tables.

## Supplementary Materials

### Supplementary Methods

#### Variant epidemic curves

We used data from CDC’s national genomic surveillance program, which aggregates biweekly SARS-CoV-2 sequence data from CDC-funded programs and public data repositories [103], to estimate COVID-19 cases attributable to variants circulating from July 2021 to September 2024. Using previously defined SARS-CoV-2 lineage groups [73], we included the following variants that attained 5% prevalence or greater anytime from July 20, 2021, to September 14, 2024: B.1.617.2, B.1.1.529, BA.1.1, BA.2, BA.2.12.1, BA.4, BA.5, BA.4.6, BF.7, BQ.1, BQ.1.1, XBB.1.5-like, XBB.1.16-like, EG.5-like, XBB, XBB.2.3, FL.1.5.1-like, HV.1, HK.3-like, JD.1.1, JN.1-like, KQ.1-like, KP.2-like, JN.1.16-like, KP.3, LB.1-like, and KP.3.1.1-like lineages. XBB and JN.1 lineages with identical spike residue 31 and receptor binding domain substitutions (residues 332–527) were grouped and denoted as “representative lineage-like.” We estimated biweekly numbers of COVID-19 infections attributable to these variants by multiplying counts of positive SARS-CoV-2 positive test results from the National Respiratory and Enteric Virus Surveillance System (NREVSS) with biweekly variant proportions from genomic surveillance [91]. For each included variant, we restricted to biweeks in which it circulated at 0.1% share or greater, and normalized to the maximum cases per variant. No additional adjustments were made for overall decreasing detection of COVID-19 cases over time due to reductions in testing availability.

Biweekly estimates of normalized variant-attributable COVID-19 cases generally appeared to be symmetrically distributed over time (Supplementary Figure 1). Periods of sequential variant takeover and predominance, such as during early Omicron evolution from B.1.1.529/BA.1 to BA.2 to BA.4/5 in 2022, appeared to have staggered variant epidemic curves (Supplementary Figure 1A). Periods of more prolonged co-circulation, such as during the latter half of 2023 when multiple XBB descendants circulated, showed greater overlap in variant epidemic curves (Supplementary Figure 1B). The peak number of attributed cases per variant occurred a median of 12 weeks after the variant first reached 0.1% prevalence. Across the variants included, the median epidemic curve standard deviation was 6.0 weeks. Assuming normally distributed time series, this indicated that approximately 95% of cases for a given variant occurred within a 24-week (5.5 month) period.

#### Variant co-circulation model

Based on our empiric assessments of variant circulation, we used a simple deterministic model to represent dynamics of two SARS-CoV-2 lineages, adapted from Bond et al.’s approach of defining symmetric epidemic curves using the binomial distribution for influenza transmission [98]. This approach allows full parametric determination of the shape and height of an epidemic curve but does not assume a mechanistic model underlying transmission.

Formally, epidemic dynamics for variants 1 and 2 are assumed to each follow a normally distributed time series 𝑃_𝑖_, with 𝑇_1_ and 𝑇_2_ representing the weeks in which variants 1 and 2 peak, respectively. Variant 1 is assumed to peak at 15 weeks, and variant 2 is assumed to peak 2 to 30 weeks after variant 1 (Figure 1); this degree of right-shift is the main variable that is adjusted to generate different variant circulation patterns. The standard deviation 𝜎 from the peak of the epidemic time series is assumed to be 6 weeks for both variants, corresponding to 95% of cases for a given variant occurring within a 24-week period.

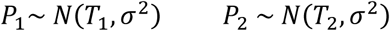

We define *U(t)* and *V(t)* as the number of unvaccinated and vaccinated individuals, respectively, at time *t*. The total population size [*N = U(t)* + *V(t)*] is fixed with the assumption of no births, deaths, or migration in or out of the population. We assume the total population size (e.g., VE study recruitment catchment area) is 1 million individuals with no age or other demographic strata modeled. The number of vaccinated individuals is represented by a piecewise linear function that increases until a pre-determined threshold 𝑉_𝑚𝑎𝑥_ at time 𝑇_𝑣_. We set 𝑉_𝑚𝑎𝑥_ to be 20% of the total population and 𝑇_𝑣_ to be 10 weeks, as updated 2023–2024 COVID-19 vaccination uptake in the US appeared to plateau at approximately 20% of the eligible adult population several months after updated doses were available [13,21,59,65].

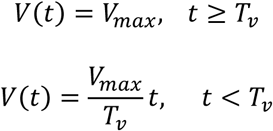

The numbers of unvaccinated and vaccinated individuals hospitalized following infection from variant 1 and recruited into the study are given by *U_1_(t)* and *V_1_(t),* respectively:

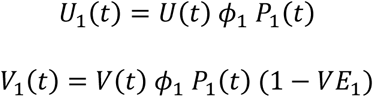

where 𝜙_1_is a parameter that scales the height of the epidemic curve (representing multiple factors not individually modeled here, including infectiousness and severity of variant 1) and *VE_1_* represents time-invariant VE specific to variant 1. We assume time-invariant VE against variants 1 and 2 (*VE_1_* = 50% and *VE_2_* = 20%, respectively), representing approximate levels of recent VE against hospitalization observed empirically against strain-matched and divergent variants, respectively [81]. Letting 𝜙_1_ = 𝜙_2_, the dynamics of variant 2 are similar:

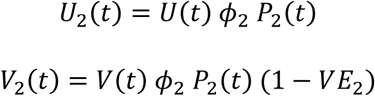

In a sensitivity analysis, we extend this model by allowing VE to change by time since vaccination 𝜏. Similar to the approach used by Lin et al. [13], we define piecewise functions such that VE increases from 0% at 𝜏 = 0 to a peak at 𝜏 = 4 weeks, representing the mounting of the immune response following vaccination; VE then subsequently wanes, representing natural waning of immunologic protection over time. Denoting 𝑉𝐸_𝑘_(𝜏) as the time-varying VE function specific to variant *k*:

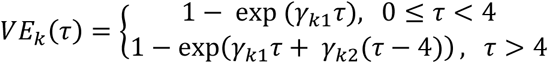

Where 𝛾_𝑖_ are parameters for adjusting the rates of increase and waning and time-varying VE functions are constrained to be greater or equal to 0. We selected 𝛾_𝑖_ such that 𝑉𝐸_1_(𝜏) peaks at 50% and 𝑉𝐸_2_(𝜏) peaks at 20% (identical to VE under the time-invariant setting), 𝑉𝐸_2_(𝜏) wanes to ∼0% by 35 weeks, and 𝑉𝐸_1_(𝜏) has a similar rate of waning (Supplementary Figure 3). These parameters represent illustrative scenarios and are not meant to be best estimates of current rates of waning of COVID-19 VE.

Under time-varying VE, the numbers of vaccinated individuals hospitalized following infection from variant *k* and recruited into the study is given by *V_k_(t)*, which combines the number of newly vaccinated individuals Δ𝑉 and the time-varying VE function for each week through a convolution:

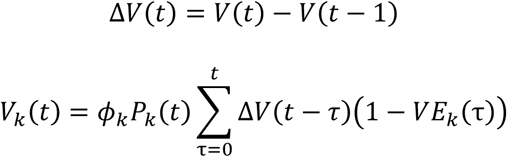

Test-negative controls can present with infection from multiple acute respiratory illness-causing pathogens and are used to estimate vaccine uptake. Numbers of enrolled test-negative control-patients are assumed to accrue at a constant rate over time. Example time series are shown in Figure 1 for counts of patients with variant 1, variant 2, and test-negative controls stratified by vaccination status.

#### Estimating vaccine effectiveness

We simulate this model for 52 weeks, which produces a time series of vaccinated and unvaccinated cases for each SARS-CoV-2 lineage that is then used as input in a logistic regression model for estimating VE. The log odds of case versus control status was modeled as a linear function of vaccination status and categorical calendar week. In simulations where VE was assumed to be time-variant, VE was calculated as 1 minus the odds ratio and compared to the defined VE value. In the setting of time-varying VE, an empirical reference VE was estimated from the full time series data, representing a weighted average of VE over the observed time since vaccination distribution. For period-based analyses with a specified threshold α, the predominance period for variant 1 was defined from the start of COVID-19 circulation (1 or more cases detected) to the last week in which variant 1’s prevalence was above α. The predominance period for variant 2 spanned from the first week in which variant 2’s prevalence was above α to the end of COVID-19 circulation. For sequence-based analyses, correct lineage determination was assumed to occur in 60% of patients, reflecting possible sample reduction from specimens with low viral load [99]. Our simplified model does not incorporate differences among patients with and without successful sequencing results.

---

**Supplementary Figure 1.**
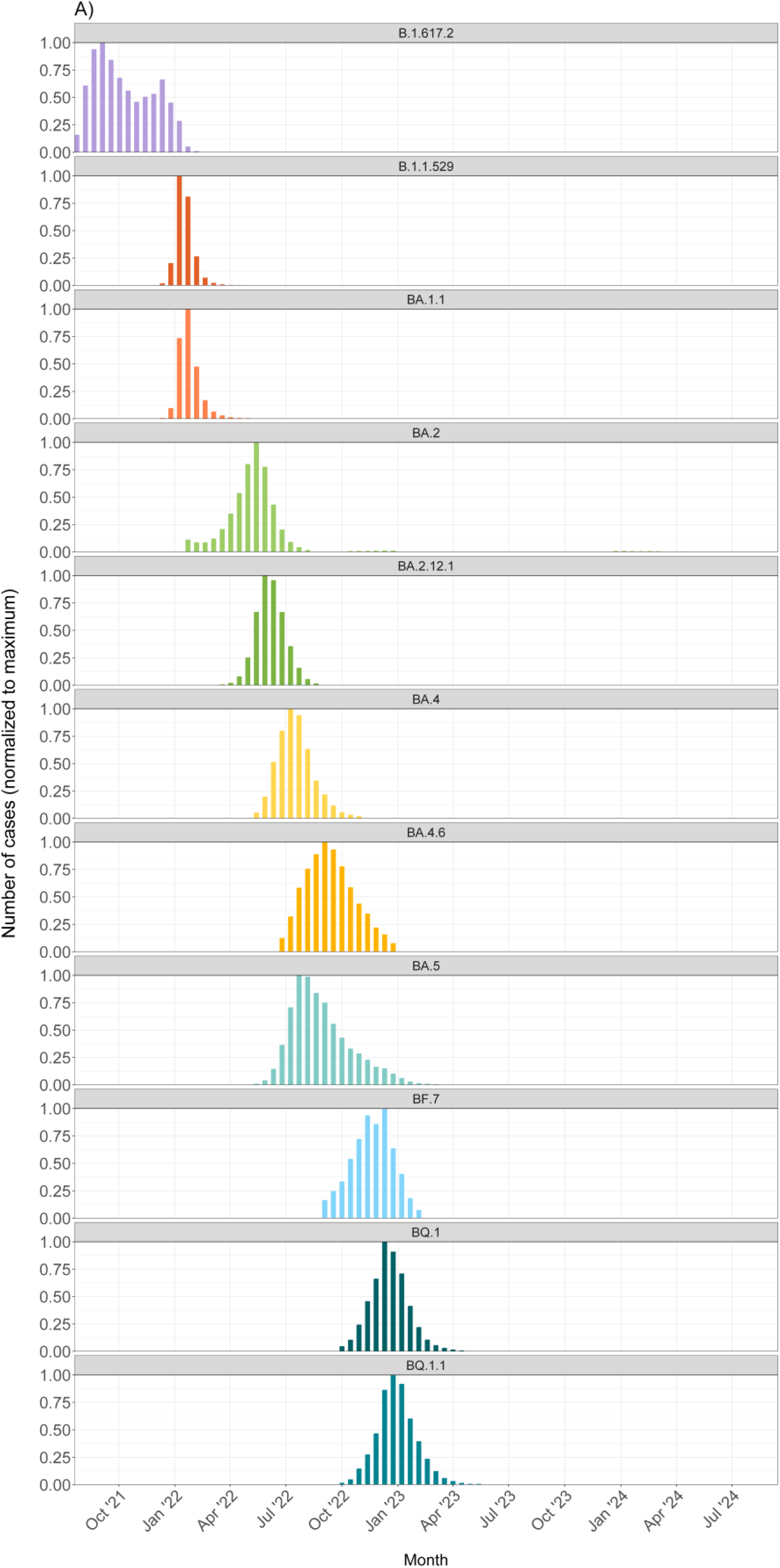
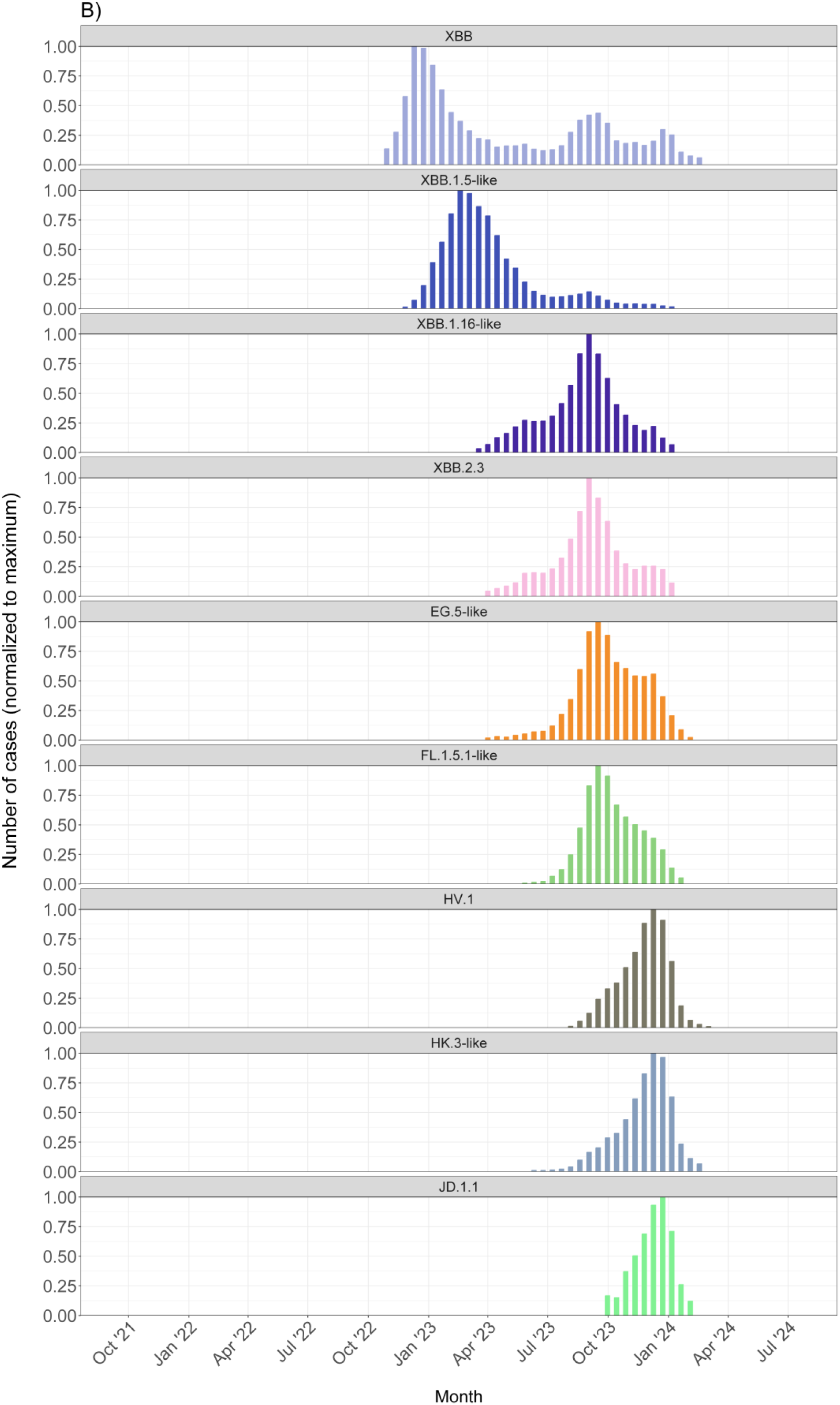
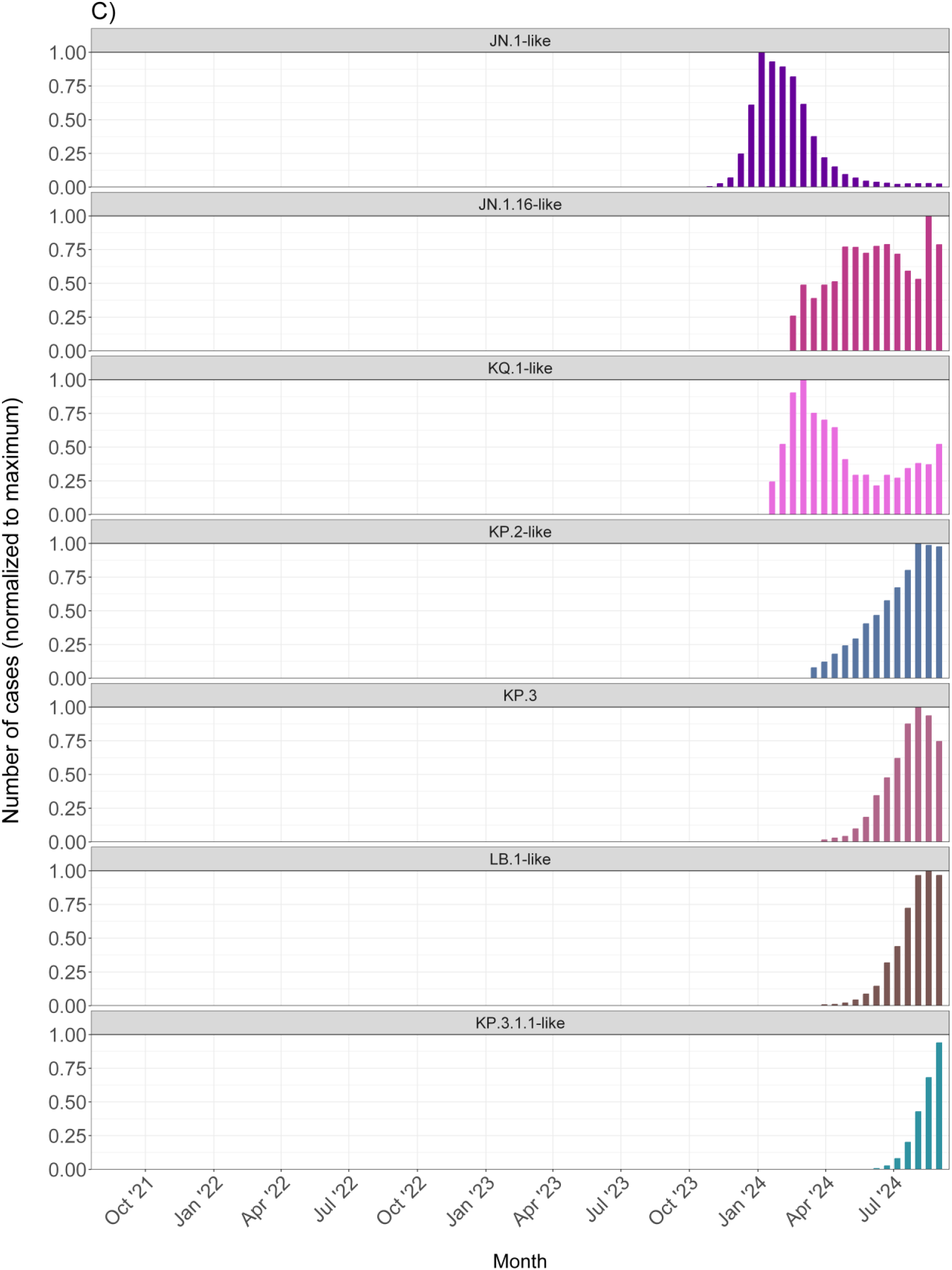
**Normalized estimated biweekly variant-attributed COVID-19 cases among A) early Omicron variants, B) XBB and descendants, and C) JN.1. and descendants** Data from CDC’s national genomic surveillance program [103] were used to estimate COVID-19 cases attributable to variants circulating from July 2021 to September 2024 among previously defined SARS-CoV-2 lineage groups [73]. We estimated biweekly numbers of COVID-19 infections attributable to these variants by multiplying counts of positive SARS-CoV-2 positive test results from the National Respiratory and Enteric Virus Surveillance System (NREVSS) with biweekly variant proportions from genomic surveillance [81]. For each variant, we restricted to biweeks in which it circulated at 0.1% share or greater and normalized to the maximum cases per variant.

**Supplementary Figure 2.**
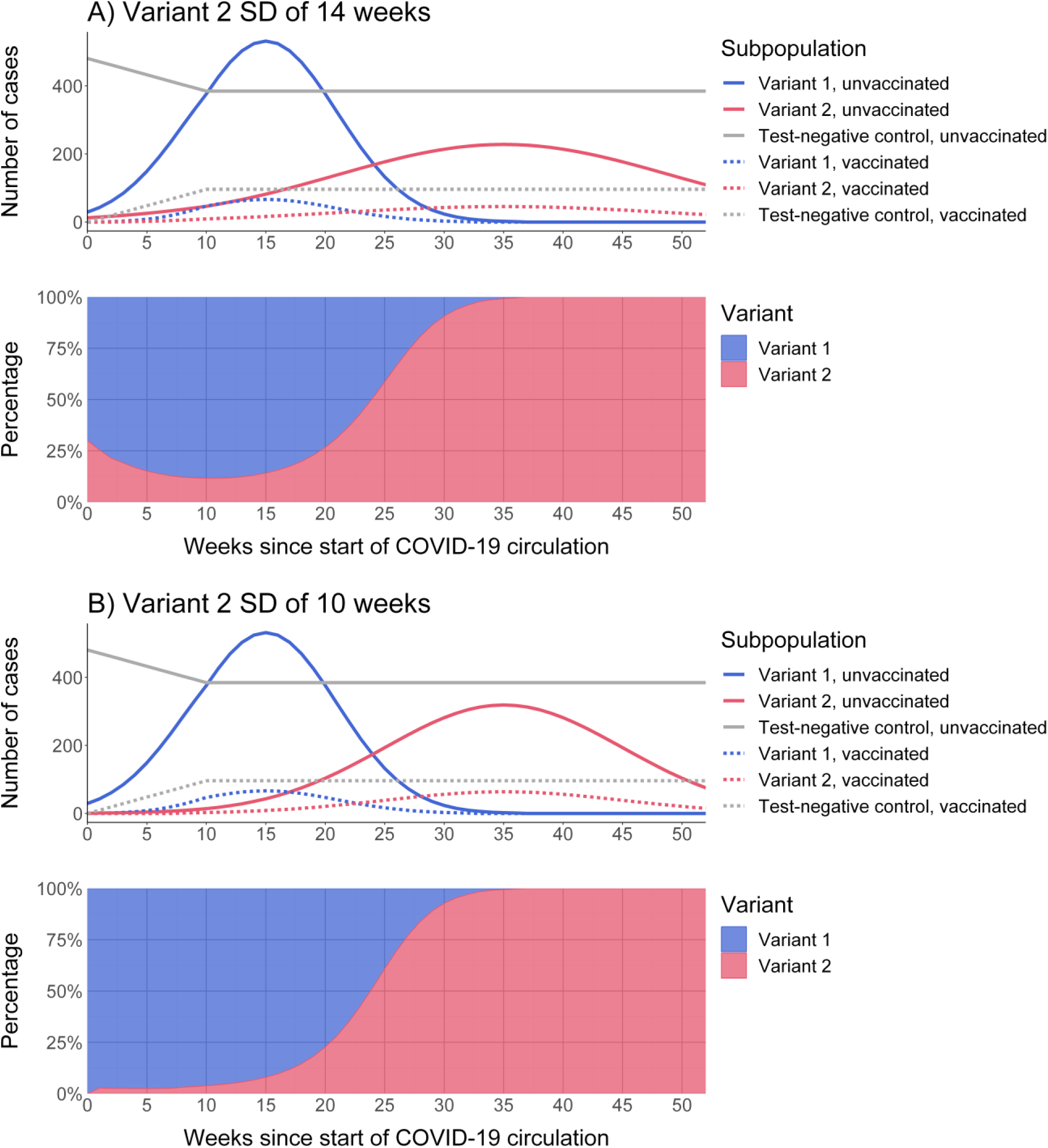
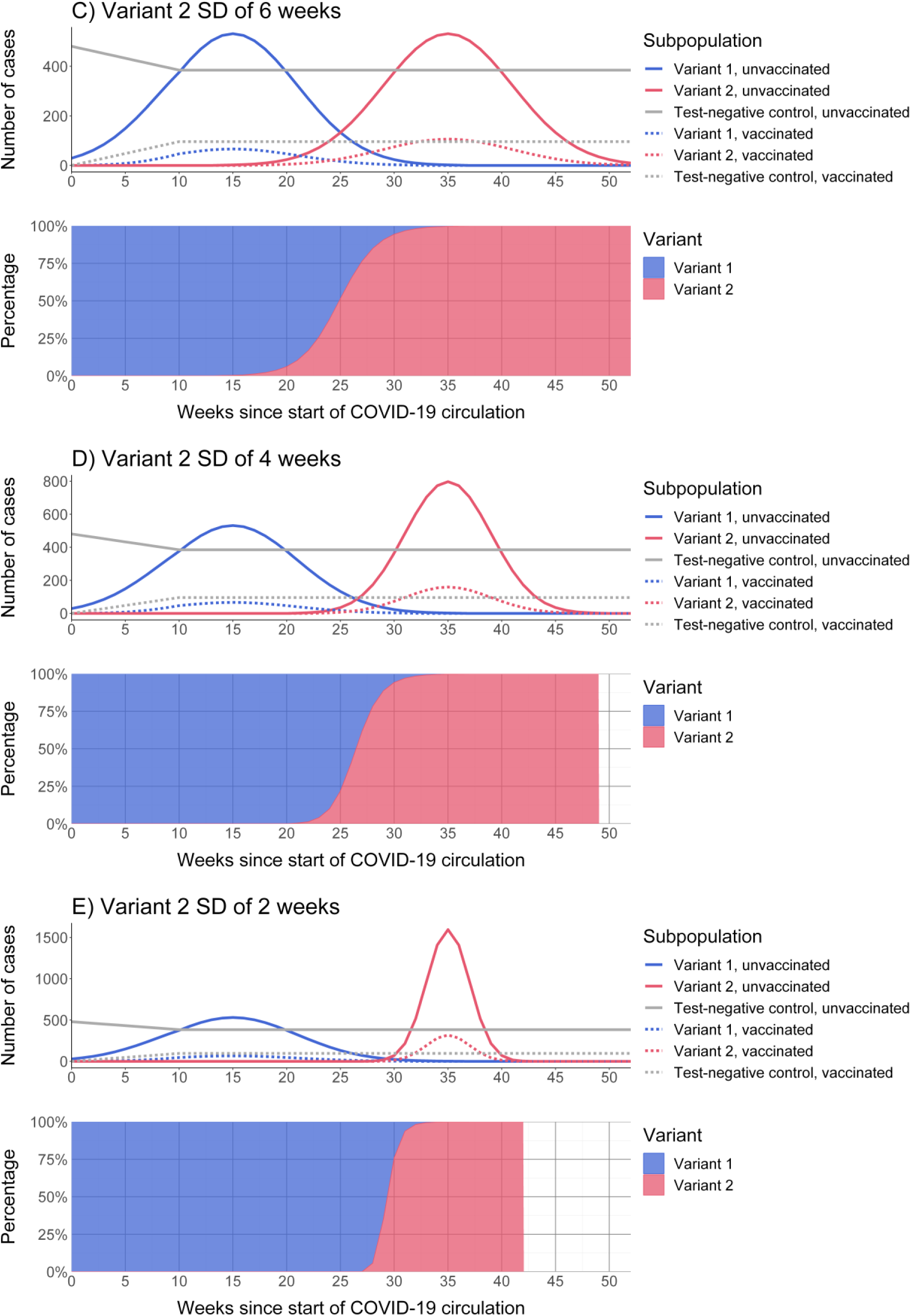
**Counts of simulated cases and controls by variant and vaccination status (top) and proportions of variants (bottom) over time for a range of standard deviations for the variant 2 epidemic curve.** Variant co-circulation was modeled using symmetric normally-distributed epidemic curves, with the standard deviation of the variant 2 epidemic curve ranging from 2 to 14 weeks. Time-invariant VE was assumed to be 50% against variant 1 and 20% against variant 2, representing approximate differences in VE observed empirically against recent strain-matched and divergent variants, respectively. Vaccine coverage was assumed to increase linearly from the start of the season before plateauing at 20%, derived from empirical trends in vaccination coverage 2023–24 and 2024–25 in the US.

**Supplementary Figure 3.**
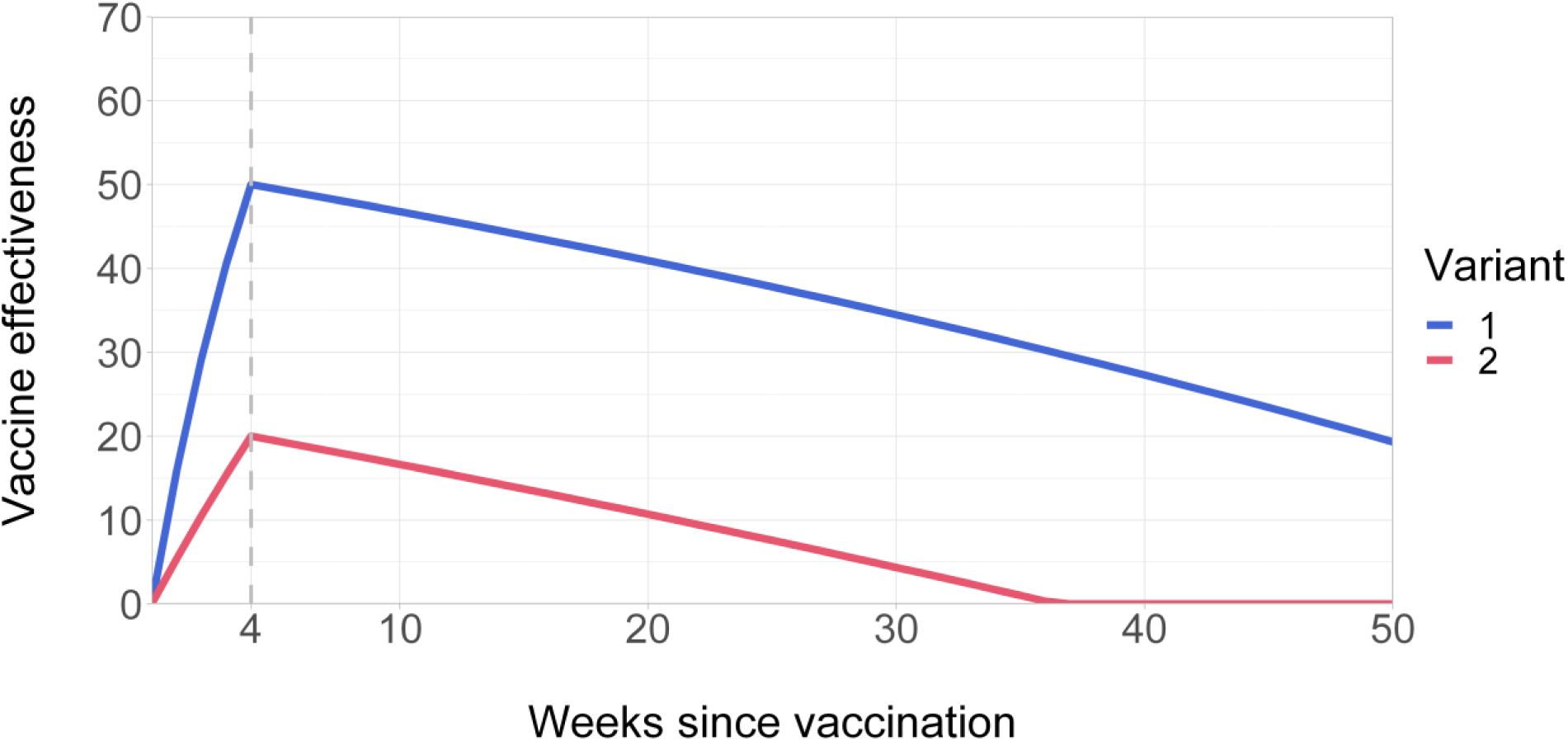
**Time-varying vaccine effectiveness by number of weeks since vaccination for variant 1 and 2.** Time-varying vaccine effectiveness was defined using the piecewise function approach of Lin et al. [81]. In this model, VE increases from 𝜏 = 0 to a peak at 𝜏 = 4 weeks of 50% against variant 1 and 20% against variant 2, representing the mounting of the immune response following vaccination; VE then subsequently wanes to 0%, representing natural waning of immunologic protection over time.

**Supplementary Figure 4.**
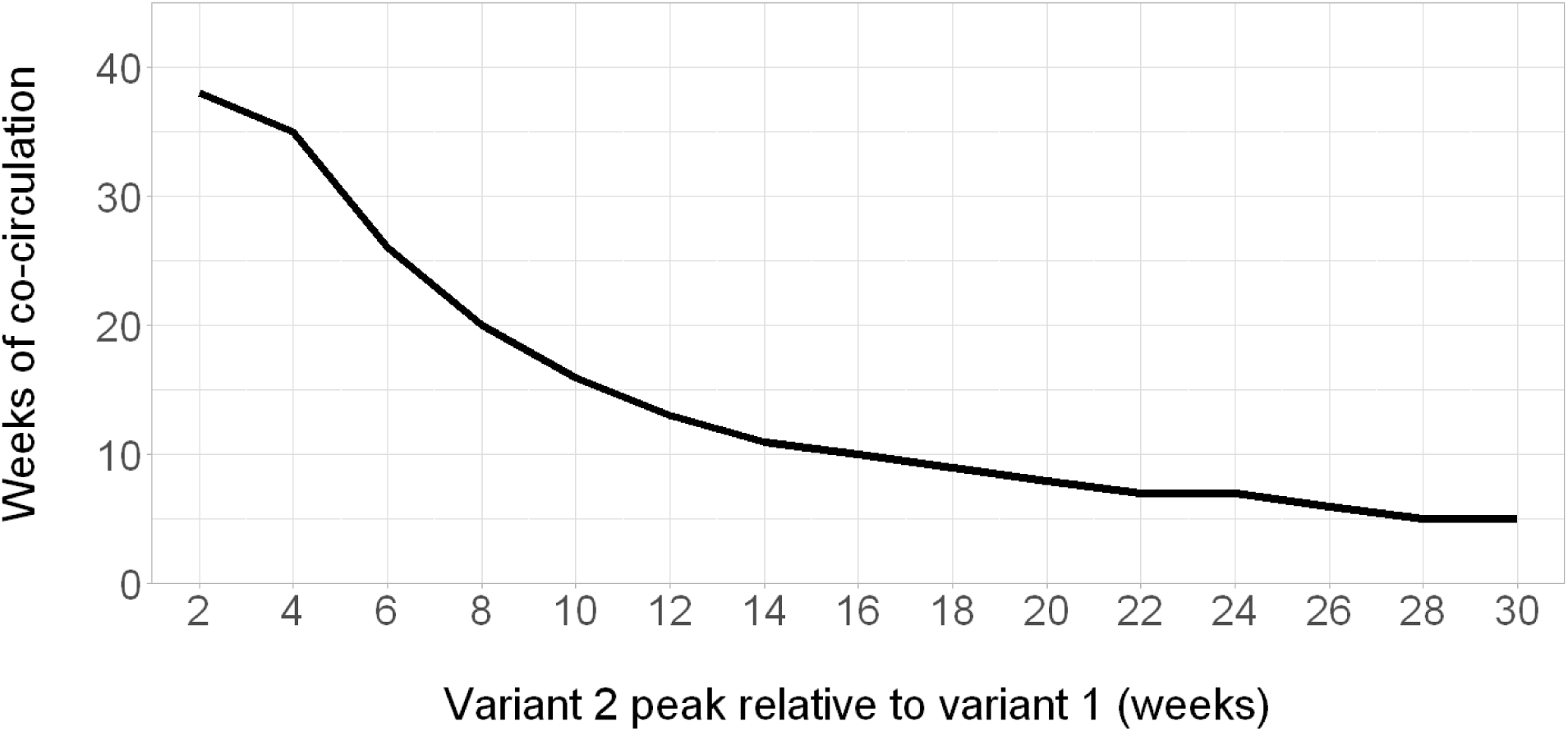
**Delay between peak of variant 1 and 2 versus duration of co-circulation in simulated epidemic curves.** Co-circulation was defined as weeks in which both variants had a prevalence of 10% or greater. Variant co-circulation was modeled using symmetric normally-distributed epidemic curves, with the peak of variant 2 circulation occurring a range of 2 to 30 weeks after variant 1.

**Supplementary Figure 5.**
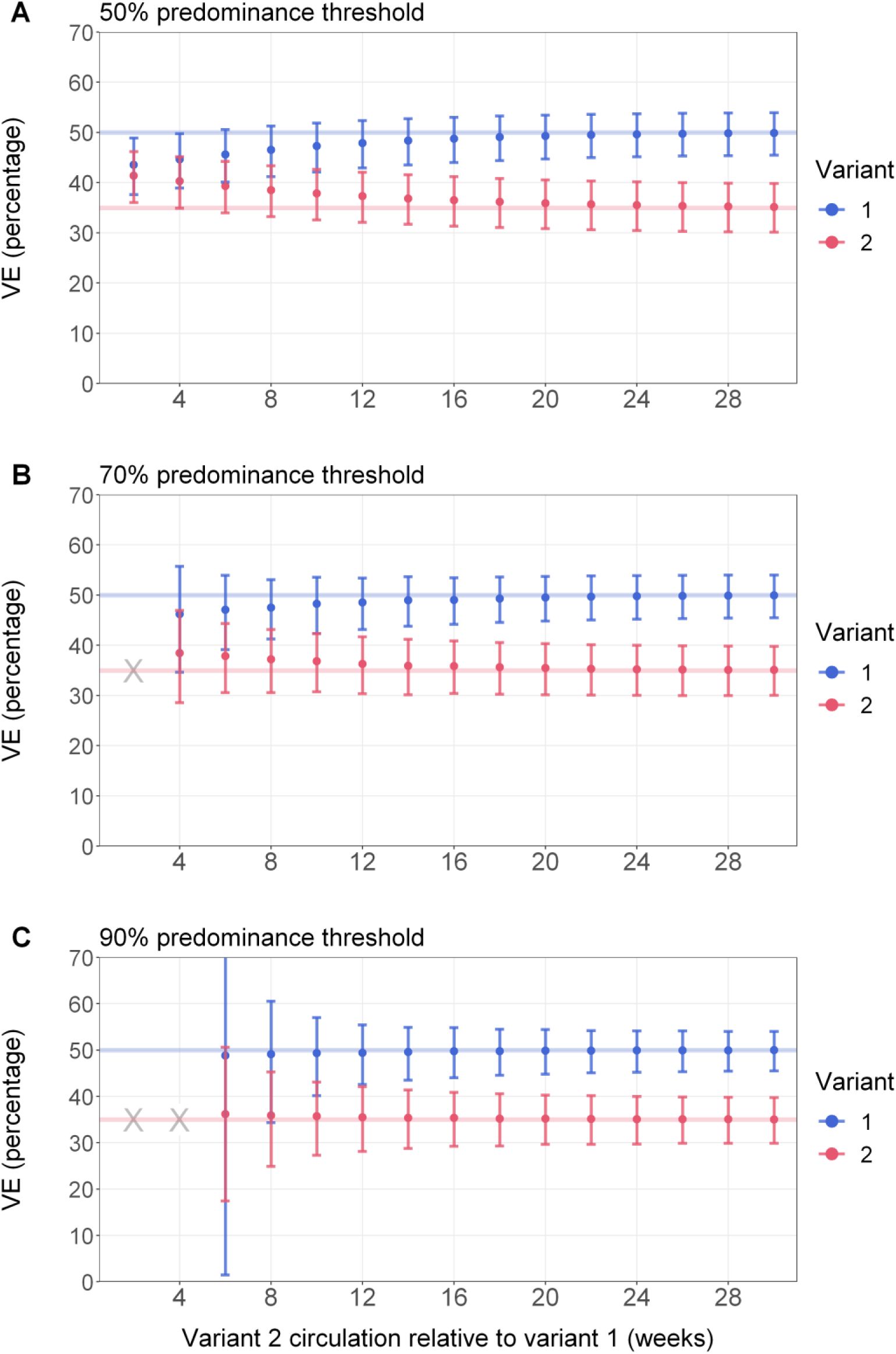
**Point estimates and 95% confidence intervals for period-based vaccine effectiveness estimates across different variant co-circulation scenarios and predominance thresholds with higher variant 2 vaccine effectiveness.** X = scenarios during which a predominance threshold was not met or the number of COVID-19 cases was <100 for either variant and VE could not be estimated. Variant co-circulation was modeled using symmetric normally-distributed epidemic curves, with the peak of variant 2 circulation occurring a range of 2 to 30 weeks after variant 1. In this sensitivity analysis, time-invariant VE was assumed to be 50% against variant 1 and 35% against variant 2. Vaccine coverage was assumed to increase linearly from the start of the season before plateauing at 20%, derived from empirical trends in vaccination coverage 2023–24 and 2024–25 in the US. Using a TND framework, we estimated lineage-specific VE using logistic regression adjusting for calendar time, and we estimated lineage-specific VE using a period-based analysis with different predominance thresholds.

**Supplementary Figure 6.**
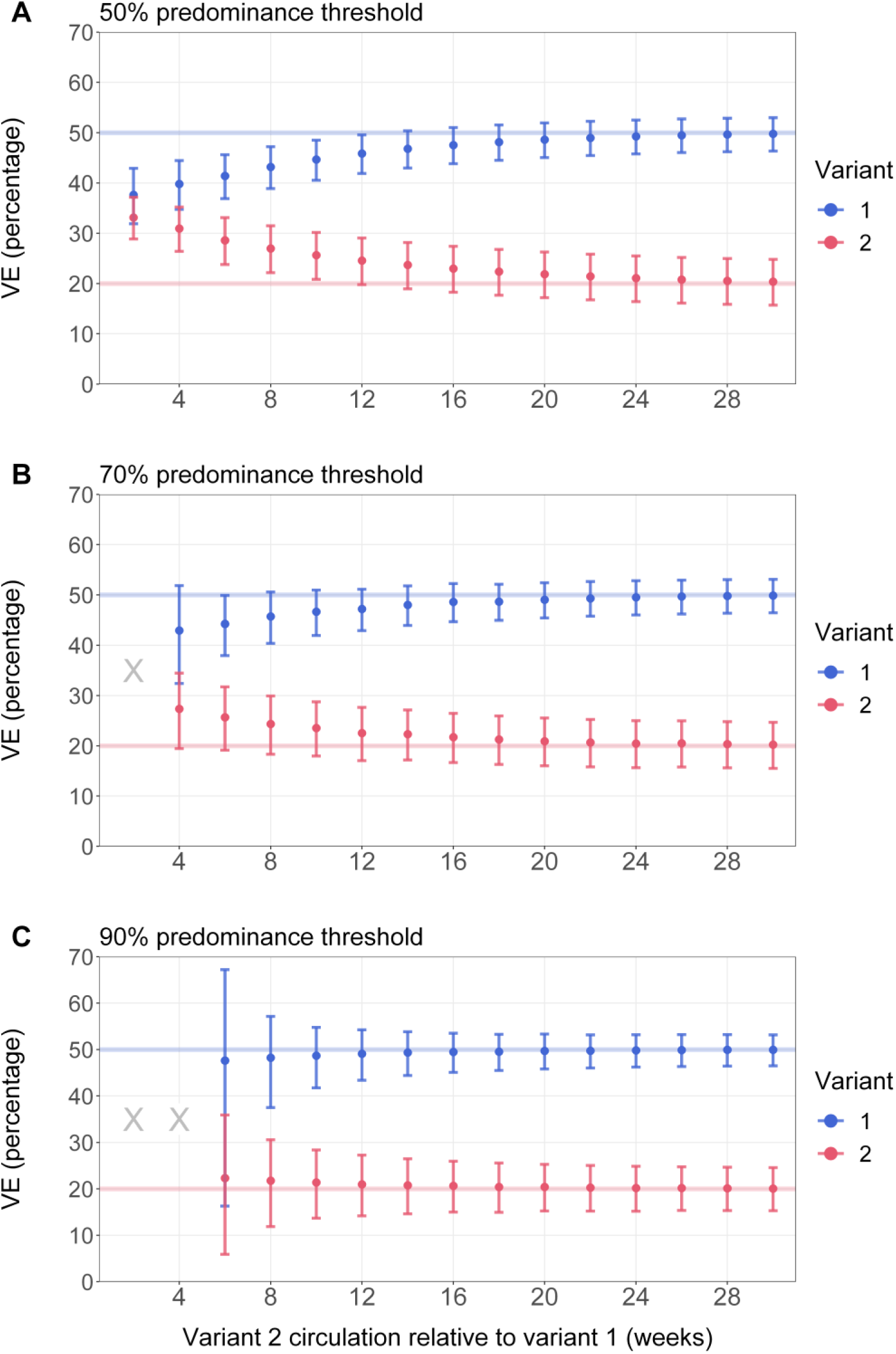
**Point estimates and 95% confidence intervals for period-based vaccine effectiveness estimates across different variant co-circulation scenarios and predominance thresholds assuming higher population-wide vaccination coverage.** X = scenarios during which a predominance threshold was not met or the number of COVID-19 cases was <100 for either variant and VE could not be estimated. Variant co-circulation was modeled using symmetric normally-distributed epidemic curves, with the peak of variant 2 circulation occurring a range of 2 to 30 weeks after variant 1. Time-invariant VE was assumed to be 50% against variant 1 and 20% against variant 2, representing approximate differences in VE observed empirically against recent strain-matched and divergent variants, respectively. In this sensitivity analysis, vaccine coverage was assumed to increase linearly from the start of the season before plateauing at 40%, potentially representing higher coverage among a subpopulation with risk factors such as older age. Using a TND framework, we estimated lineage-specific VE using logistic regression adjusting for calendar time, and we estimated lineage-specific VE using a period-based analysis with different predominance thresholds.

**Supplementary Figure 7.**
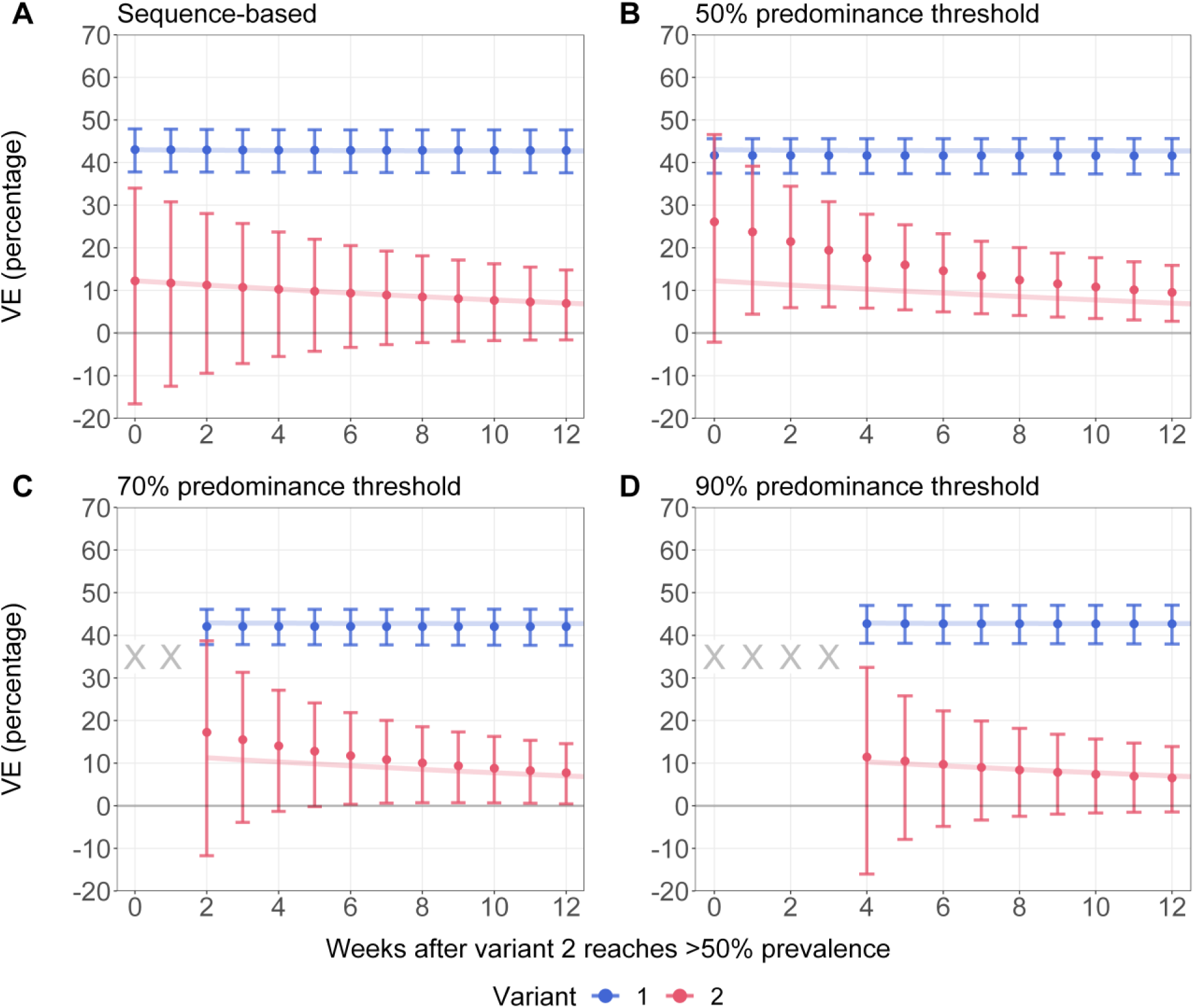
**Point estimates and 95% confidence intervals for (A) sequence-based and (B-D) period-based vaccine effectiveness estimates across different predominance thresholds by week after variant 2 reaches 50% prevalence allowing for time-varying vaccine effectiveness.** X = scenarios during which a predominance threshold was not met or the number of COVID-19 cases <100 for either variant and VE could not be estimated. Variant co-circulation was modeled using symmetric normally-distributed epidemic curves, with the peak of variant 2 circulation occurring a range of 2 to 30 weeks after variant 1. Time-varying vaccine effectiveness was defined using the piecewise function approach of Lin et al. [80]. In this model, VE increases from 𝜏 = 0 to a peak at 𝜏 = 4 weeks of 50% against variant 1 and 20% against variant 2, representing the mounting of the immune response following vaccination; VE then subsequently wanes to 0%, representing natural waning of immunologic protection over time (Supplementary Figure 3). Vaccine coverage was assumed to increase linearly from the start of the season before plateauing at 20%, derived from empirical trends in vaccination coverage 2023–24 and 2024–25 in the US. Using a TND framework, we estimated lineage-specific VE using logistic regression adjusting for calendar time, and we estimated lineage-specific VE using a period-based analysis with different predominance thresholds.

**Table 1.** Methods, advantages, and considerations for estimating lineage-specific COVID-19 VE using different lineage assignment methods.

|  | Sequence-based | Proxy-based | Period-based |
| --- | --- | --- | --- |
| Method for lineage assignment | Whole-genome sequencing of SARS-CoV-2 positive cases | Use of a proxy genetic marker (such as spike $\Delta 69-70$ correlating with S-gene PCR target presence or failure) for a lineage of interest | Lineage assignment based on variant predominance time periods from internal or external data sources, such as national genomic surveillance or public sequence data repositories |
| Requirements and costs | Requires funding and capacity for whole-genome sequencing, including sample collection and storage; most expensive approach and sample sizes could be reduced by cost constraints | Less costly compared to whole-genome sequencing and can take advantage of existing clinical laboratory testing infrastructure | If external data sources are used, does not require direct funding or capacity for whole-genome sequencing of specimens, but still dependent on representativeness and timeliness of external sources |
| Advantages | Full genomic information allows identification of sublineages and mutations of interest even during periods of prolonged variant co-circulation | Compared to sequence-based analyses, less costly, higher throughput, and faster, and retains accuracy for categorizing broad variant groups compared to whole-genome sequencing data [30]. May have higher sample sizes compared to sequence-based analyses | Does not require collection or sequencing of specimens, enabling study platforms without laboratory capacity to also generate lineage-specific estimates. May have higher sample sizes compared to sequence-based analyses |
| Disadvantages and potential biases | Specimens that cannot be sequenced due to viral load may be dropped out; sensitivity analyses can be conducted | Cannot resolve specific spike protein substitutions of interest or sublineage evolution | Subject to lineage misclassification bias which could result in underestimation of the difference in VE by lineage |
|  | comparing overall VE with VE among sequenced patients to assess degree of bias [28,29]<br>Sequencing data lags need to be minimized for timely VE estimation to inform public health decision-making | Requires continued validation that proxies correlate well with lineage categories identified using whole-genome sequencing | Requires setting a threshold to define predominance, which has varied in the literature (50–90%) and could affect the extent of misclassification<br>Cannot estimate lineage-specific VE during prolonged periods with multiple co-circulating variants<br>Can be challenging to disentangle effect of variants from other time-varying confounders |
| Analytic opportunities | Leveraging the full extent of genomic information available to assess VE by spike protein substitutions or genetic distance [68] | Characterization of the impact of proxy misclassification error and how model-based corrections can be used to improve estimates | Development of new analytic approaches to estimate VE during periods of prolonged co-circulation, including incorporating empirical variant proportions as a covariate in regression models [82] or imputing infecting variant probabilistically |

**Table 2.** Parameters used in lineage vaccine effectiveness simulations.

| Parameter | Value or distribution | Rationale and references |
| --- | --- | --- |
| Population size (N) | 1,000,000 |  |
| Duration | 52 weeks |  |
| Vaccination coverage | Linear increase from 0% to 20% by 10 weeks | Derived from empirical trends in vaccination coverage 2023–24 and 2024–25 in the US [13,21,59,65] |
| Time-invariant lineage-specific VE against hospitalization ($VE_1$ and $VE_2$) | $VE_1 = 50\%$<br>$VE_2 = 20\%$ | Derived from VE against XBB and JN.1 lineage hospitalization during administration of 2023–24 COVID-19 XBB.1.5 vaccines [81] |
| **Time-varying lineage-specific VE** against hospitalization ($VE_1(\tau)$ and $VE_2(\tau)$) | VE starts at 0% and peaks 4 weeks after vaccination at 50% and 20% against variant 1 and 2, respectively; VE wanes after peaking (Supplementary Figure 3) | Waning vaccine effectiveness was defined using a piecewise function approach adapted from Lin et al. [91] (Supplementary Methods) |
| Variant epidemic time series | $P_1 \sim N(T_1, \sigma^2)$<br>$P_2 \sim N(T_2, \sigma^2)$ | Model framework adapted from Bond et al. [98] |
| Incidence of non-SARS-CoV-2 acute respiratory illness pathogens | Uniform | Numbers of enrolled test-negative control-patients are assumed to accrue at a constant rate over time |
| Variant epidemic time series standard deviation ($\sigma$) | 6 weeks | Derived from empirical trends in variant circulation (Supplementary Methods). Corresponds to 95% of cases for a given variant occurring within a 24-week period |
| Variant 1 peak ($T_1$) | 15 weeks | |
| Variant 2 peak ($T_2$) | 17 to 45 weeks (2 to 30 weeks after variant 1 peak) | Range of values was selected to generate variety in co-circulation patterns |
| Percent of cases sequenced in sequence-based analysis | 60% | Derived from empirical data from a multisite inpatient TND network [10,19,54–57] |
| Predominance thresholds $\alpha$ used in period-based analyses | 50%, 70%, 90% | Range of values was selected to include thresholds used in previous analyses, such as 50% [11,58–60], 60% [61], 70% [62], 75% [12,63–65], 80% [66], and 90% [99] |

## Supplementary Tables

Supplementary Table 1. Simulation results underlying Figure 2 (Point estimates and 95% confidence intervals for period-based weekly vaccine effectiveness estimates across different variant co-circulation scenarios and predominance thresholds).

Supplementary Table 2. Simulation results underlying Figure 3 (Point estimates and 95% confidence intervals for sequence-based and period-based weekly vaccine effectiveness estimates across different predominance thresholds by week after variant 2 reaches 50% prevalence).

## Notes

### Funding Statement

This study did not receive any external funding.

### Author Declarations

The study used openly available data located at https://data.cdc.gov/Laboratory-Surveillance/SARS-CoV-2-Variant-Proportions/jr58-6ysp/about_data and https://data.cdc.gov/Laboratory-Surveillance/Percent-Positivity-of-COVID-19-Nucleic-Acid-Amplif/gvsb-yw6g/about_data

### Summary of Updates

This version incorporates revisions made in response to peer review.

